# A Comparative Analysis of COVID-19 Physical Distancing Policies in Botswana, India, Jamaica, Mozambique, Namibia, Ukraine, and the United States

**DOI:** 10.1101/2021.02.09.21251433

**Authors:** Jeff Lane, Arianna Means, Kevin Bardosh, Anna Shapoval, Ferruccio Vio, Clive Anderson, Anya Cushnie, Norbert Forster, Jenny Ledikwe, Gabrielle O’Malley, Shreshth Mawandia, Anwar Parvez, Lucy Perrone, Florindo Mudender

## Abstract

**Background:** Understanding the differences in timing and composition of physical distancing policies is important to evaluate the early global response to COVID-19. A physical distancing intensity framework comprising 16 domains was recently published to compare physical distancing approaches between U.S. States. We applied this framework to a diverse set of low and middle-income countries (LMICs) (Botswana, India, Jamaica, Mozambique, Namibia, and Ukraine) to test the appropriateness of this framework in the global context and to compare the policy responses in this set of LMICs and with a sample of U.S. States during the first 100-days of the epidemic.

**Results:** All six of the LMICs in our sample adopted wide ranging physical distancing policies. The highest peak daily physical distancing intensity in each country was: Botswana (4.60); India (4.40); Ukraine (4.40); Namibia (4.20); and Jamaica (3.80). The number of days each country stayed at peak intensity ranged from 12-days (Jamaica) to more than 67-days (Mozambique). We found some key similarities and differences, including substantial differences in whether and how countries expressly required certain groups to stay at home. We also found that the LMICs generally implemented physical distancing policies when there were few confirmed cases and the easing of physical distancing policies did not discernably correlate with change in COVID-19 incidence. The physical distancing responses in the LMIC sample were generally more intense than in a sample of U.S. States, but results vary depending on the U.S. State. For example, California had a peak intensity of 4.29, which would place California below the peak intensity for Botswana, India, and Ukraine but above Mozambique, Namibia and Jamaica. The U.S. State of Georgia had a peak intensity of 3.07, which would place it lower than all of the LMICs in this sample. The peak intensity for the U.S. 12-state average was 3.84, which would place it lower than every LMIC in this sample except Jamaica.

**Conclusion:** This analysis helps to highlight the differing paths taken by the countries in this sample and may provide lessons to other countries regarding options for structuring physical distancing policies in response to COVID-19 and future outbreaks.

## Background

On 30 January 2020, the World Health Organization (WHO) declared COVID-19 a Public Health Emergency of International Concern and six weeks later on 11 March declared COVID-19 a pandemic.(1) In the weeks preceding the declaration, multiple countries proactively implemented various levels of mass physical distancing combined with other measures to interrupt COVID-19 transmission. The first mass physical distancing policy was implemented in Wuhan, China on January 23, 2020.(2) On March 13, the Director General of the WHO delivered a press briefing in which he stated “The experience of China, the Republic of Korea, Singapore and others clearly demonstrates that aggressive testing and contact tracing, combined with social distancing measures and community mobilization, can prevent infections and save lives.”(3) By the end of March, dozens of countries around the world had implemented mass physical distancing as part of a broader policy response to COVID-19, which has been referred to as “the great pause.”

Understanding the differences in the timing and composition of physical distancing policies, as they were implemented in different countries in the first half of 2020, is an important first step in efforts to evaluate the early global pandemic response to COVID. This nuanced understanding may also help guide future COVID-19 response partners identify the optimal package (or set of packages, depending on the epidemiological circumstance) of physical distancing approaches that maximizes public health benefit while minimizing social and economic harm. A recently published Social Distancing Policy Intensity Coding Framework addresses policy specification by organizing social distancing policies according to 16 physical distancing policy domains.(4) The framework was used to describe the physical distancing policy responses in 12 U.S. States. This framework has not been used to analyze physical distancing policy approaches in low and middle-income countries (LMICs). Therefore, its appropriateness for the LMIC context remains unknown.

To test the appropriateness of the above framework and to facilitate comparison of physical distancing approaches in a diverse set of LMICs, we applied the Social Distancing Policy Intensity Framework (referred to as the Physical Distancing Intensity Framework) to describe and compare the COVID-19 physical distancing policy responses in six LMICs: Botswana, India, Jamaica, Mozambique, Namibia, and Ukraine. We analyzed the policy approaches during the first 100 days following the WHO declaration of COVID-19 as a pandemic (through June 19, 2020). We compare the range and temporal dimensions of these physical distancing policy responses in these six LMICs. We also reflect on the similarities and differences between physical distancing policies in the United States (U.S.) and LMIC settings.

## Methods

The University of Washington’s International Training and Education Center for Health (UW/I-TECH) operates global health programs in more than a dozen countries and established an incident command structure to monitor the policy responses to COVID-19 in countries where I-TECH works. This analysis focuses on six LMICs with active UW/I-TECH programs: Botswana, India, Jamaica, Mozambique, Namibia, and Ukraine. Demographic and other attributes that may affect country policy making in response to the COVID-19 pandemic are summarized in Table 1.

**Table 1.** Overview of country population, demographic, and health system attributes

| | Total population (million, 2016) | Life Expectancy at birth (m/f) (years, 2016) | Gross national income per capita (PPP international \$, 2013) | Total expenditure on health per capita (Intl \$, 2014) |
| --- | --- | --- | --- | --- |
| Botswana(5) | 2.25 | 64/68 | 15,500 | 871 |
| India(6) | 1,324.17 | 67/70 | 5,350 | 267 |
| Jamaica(7) | 2.88 | 74/78 | 8,480 | 476 |
| Mozambique(8) | 28.83 | 58/62 | 1,040 | 79 |
| Namibia(9) | 2.48 | 61/66 | 9,590 | 869 |
| Ukraine(10) | 44.44 | 68/77 | 8,960 | 584 |
| United States(11) | 322.18 | 76/81 | 53,960 | 9,403 |

Our analysis draws upon the Physical Distancing Intensity Framework, initially developed to analyze policy responses in 12 U.S. states.(4) This framework consists of 16 physical distancing policy domains that are scored using an ordinal intensity scale of 0-5, including: social gatherings; religious gatherings; funerals; stay at home orders; restaurants; bars; movie theatres; hair salons and barbers; indoor gyms; non-essential retail stores; childcare; K-12 schools; higher education; nursing homes; prisons; and voting. Low intensity scores indicate the absence of lockdown policies or policies that are minimally restrictive, while higher intensity scores indicate lockdown mandates that are more comprehensive and restrictive.

We made minor adaptations to the Physical Distancing Intensity Framework for this analysis. We removed the “voting” and “nursing home” domains since they were not relevant for the LMIC sample (no national elections took place in the March-June 2020 timeframe, and only 2 of 6 LMICs (Namibia and Jamaica) had policies involving nursing homes). We also added one domain (public transportation) that was not included in the initial U.S. Physical Distancing Intensity Framework but was common across LMIC policies we reviewed. Thus, 15 domains were included in this analysis. Each domain was scored using an ordinal intensity scale of 0-5 (Scale: 0 = no mandate or recommendations; 1 = recommendations only; 2 = mandate-low intensity; 3 = mandate-medium intensity; 4 = mandate-high intensity; 5 = mandate-very high intensity).

National level policy documents were collected from public government websites by team members based in each country. National policies adopted between January 1, 2020 and June 19, 2020 (100 days after the WHO declared COVID-19 to be a pandemic) were included in the analysis. Daily physical distancing policies were coded longitudinally using the Physical Distancing Intensity Framework. Policies from Botswana, India, Jamaica, and Namibia, which were available in English, were reviewed and initially coded by one co-author (JL). Policies from Ukraine, which were only available in Ukrainian, were reviewed and coded by another co-author (AS), who is fluent in Ukrainian and English. Policies from Mozambique, which were only available in Portuguese, were reviewed and coded by two co-authors (FM and FF), who are both fluent in Portuguese and English. Excel was used to capture the results of the coding described above. The final coding was reviewed by at least one co-author residing in each sample country. An average daily intensity score was calculated for each country by summing the domain specific intensity on that day and dividing by the total number of domains (i.e., 14 or 15 domains). Policy data for the 12 U.S. State sample were obtained from Lane, et al. and is publicly available.(12) The coded policy intensity data for all sample countries is available as supplementary information.

To descriptively characterize the relationship between national physical distancing policies and country-specific epidemic curves, we collected COVID-19 confirmed case and mortality data for the sample countries from the Johns Hopkins University COVID-19 dataset, over the same period of time.(13) Policy and prevalence data are presented side by side to identify patterns in the timing of policies in relation to documented case counts.

These trajectories were compared to daily policy intensity in the U.S. The U.S. physical distancing policy response was completely state-based, so we used to the U.S. states of California and Georgia to illustrate the variability in the USA state-based response. California had the highest peak intensity of the 12-US state sample (tied with Colorado, but California peaked earlier than Colorado). Georgia had the lowest peak average daily intensity from the U.S. 12-state sample. We also calculated an average daily intensity across the 12-state sample using the 14 domains common to the U.S. analysis and this analysis of LMICs.

This analysis relied solely on publicly available data sources and therefore no human subjects research review was required.

## Results

All six of the LMICs in our sample adopted wide ranging physical distancing policies such as closing schools, restricting the occupancy of certain commercial locations, and limiting the size of gatherings. Each country adopted policies mandating some form of physical distancing in at least 14 out of 15 domains in our framework. Four countries (Botswana, Jamaica, Namibia, and Ukraine) mandated some level of physical distancing in all 15 domains. Two countries (India and Mozambique) imposed physical distancing in 14 out of 15 domains. Table 2 shows the date of the first mandate in each country, the peak policy intensity, and the time span for peak intensity.

**Table 2.** Physical distancing intensity by country (15 domains)

| Jurisdiction | Date of first policy mandating physical distancing (2020) | Peak daily policy intensity* | Number of days at peak intensity | Date range of peak intensity (2020) |
| --- | --- | --- | --- | --- |
| Botswana | March 16 | 4.60 | 35 days | April 2 – May 7 |
| India | March 16 | 4.40 | 46 days | April 15 – May 31 |
| Jamaica | March 25 | 3.80 | 42 days | April 8-May 18 |
| Mozambique | March 15 | 3.87 | 67+ days | April 13- (still in effect as of June 19) |
| Namibia | March 28 | 4.20 | 14 days | April 21 – May 4 |
| Ukraine | March 12 | 4.40 | 35 days | April 6 – May 10 |

\*Scale: 0 = No Mandate or Recommendations; 1 = Recommendations Only; 2 = Mandate-Low; 3 = Mandate-Medium; 4 = Mandate-High; 5 = Mandate-Very High

The country with the highest average daily intensity peak was Botswana (4.60). India and Ukraine tied for the next highest peak intensity of 4.40. Namibia had the next highest peak intensity of 4.20. Mozambique had the next highest peak at 3.87, followed by Jamaica at 3.80. The number of days each country stayed at peak physical distancing intensity ranged from 12 days (Jamaica) to more than 67 days (Mozambique).

### Physical distancing intensity by domain

As reflected in Figure 1, there were a number of similarities in peak physical distancing policie adopted amongst sampled LMICs. All countries in the sample closed K-12 schools and banned gatherings of 10 or more. Some countries, such as Botswana and India, banned even smaller gatherings. All countries adopted policies imposing restrictions on restaurants and most banned on-premises dining, with the exception of Mozambique. All countries ordered closure of bars, indoor movie theatres, and indoor gyms. Most countries banned non-essential visits to prisons, although we did not identify national policies expressly prohibiting non-essential prison visitors in India or Jamaica. Some countries, such as Mozambique, also authorized early release of prisoners to decongest prisons.(14)

**Figure 1.**
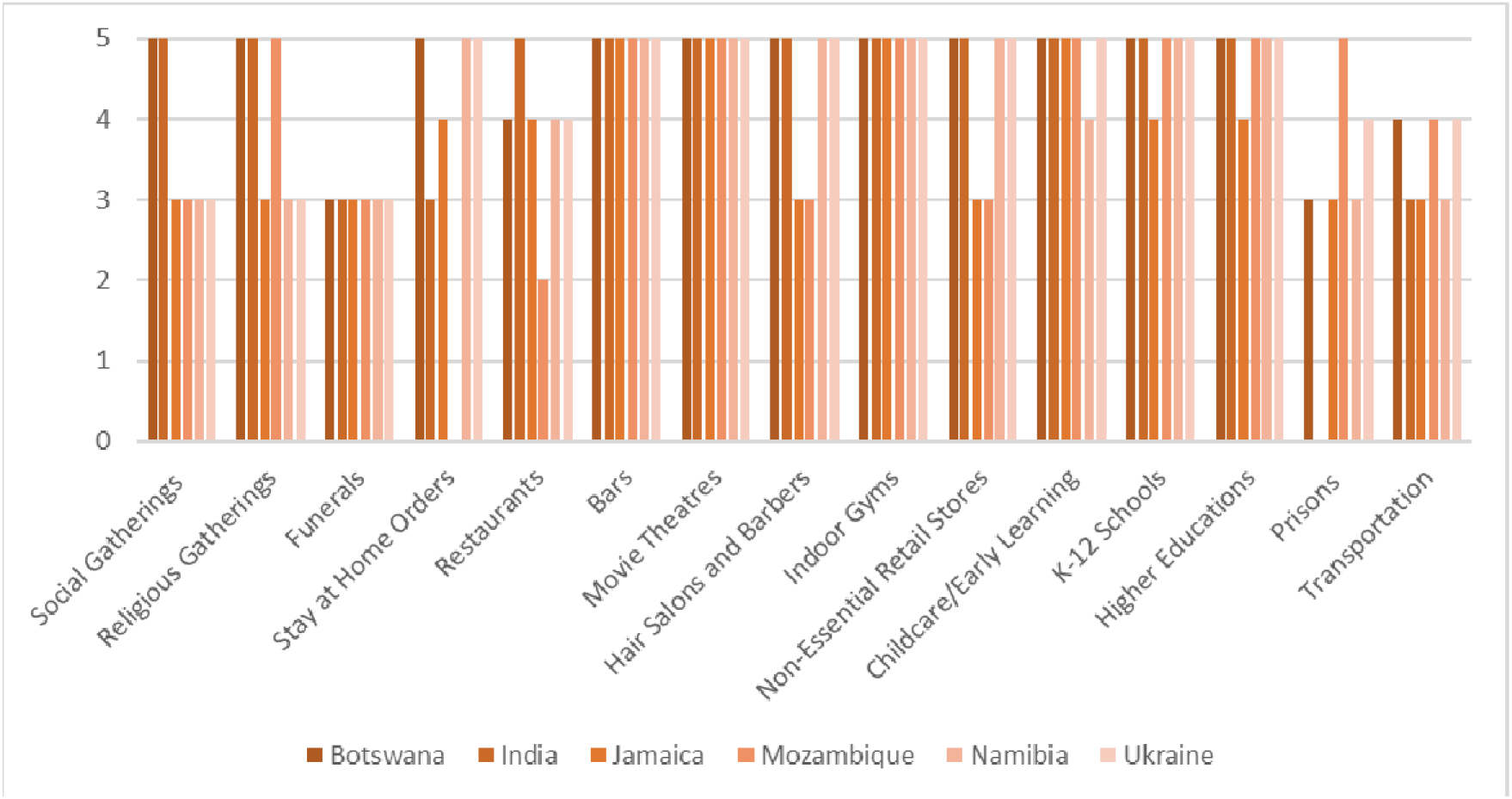
Peak intensity by domain and country

### Stay at home policies

We found substantial differences in whether and how the sampled LMICs adopted policies expressly requiring certain groups to stay at home. For example, Mozambique’s physical distancing policies did not include any explicit stay at home requirement. India and Jamaica each adopted limited stay at home orders that only applied to certain groups and imposed a curfew for all others. For example, India adopted a stay at home order limited to children under 10, students, and the elderly. Jamaica adopted a stay at home order limited to people over 75, which was later modified to apply to people over the age of 65. The stay at home policies in India and Jamaica were coded as medium intensity. Botswana, Namibia, and Ukraine each adopted intense forms of stay at home orders that required all groups to stay at home except when engaging in essential activities and included additional requirements or restrictions, such as compelling essential workers carry permission letters from their employer or limiting the number of people that can leave the household at a time. We coded the stay at home policies in Botswana, Namibia, and Ukraine as very high intensity.

### Public transportation

We observed a number of policies in this LMIC sample aimed at imposing physical distancing on public transportation, which was not included as a domain in the original US-based Physical Distancing Intensity Framework. For example, India adopted a ban on taxis, rickshaws, buses and rail service for public transport. In Mozambique, an initial policy was adopted that banned use of motorcycles, bicycles and automobile taxi services and limited buses and mini-buses to one-third occupancy. This initial approach led to protests in multiple regions of the country and the policy was relaxed approximately 1-week later.(15) Ukraine adopted an occupancy cap on public transport and limited transport to only persons carrying essential worker permits. Jamaica, Namibia, and Botswana limited hours of operation and occupancy for public transportation and Botswana further limited access to those engaging in essential activities.

### Timing of physical distancing policies

We found some key similarities and differences in the timing of physical distancing mandates. There was a wide range in the number of policy domains that were included in each country’s first physical distancing policy. For example, only two domains were covered in the first mandate issued by Mozambique on March 15. In contrast, the first physical distancing policy mandate in Namibia covered 14 domains. The domains most frequently included in the first mandate were social gatherings and religious gatherings, which were included in the first policy mandate in five out of six countries. The second most common domains included in the first policy mandate were indoor gyms, K-12 schools, childcare/early learning, and higher education, which were each included in the first policy mandate in four out of six countries. The domain least likely to be included in the first mandate were prisons (no countries).

Figure 2 demonstrates the intensity of physical distancing in Botswana longitudinally by domain, which peaked with the highest average intensity on a given day (4.60, from April 2^nd^ - May 7^th^ 2020) (Figure 2). Figure 2 illustrates that while the intensity of some domains moved in unison, the intensity of some domains diverged. For example, in Botswana two groupings of domains followed the same intensity paths for the entire time period: (1) social gatherings and religious gatherings; and (2) hair salons/barbers and non-essential retail. All other domains diverged at some point during the time period.

**Figure 2.**
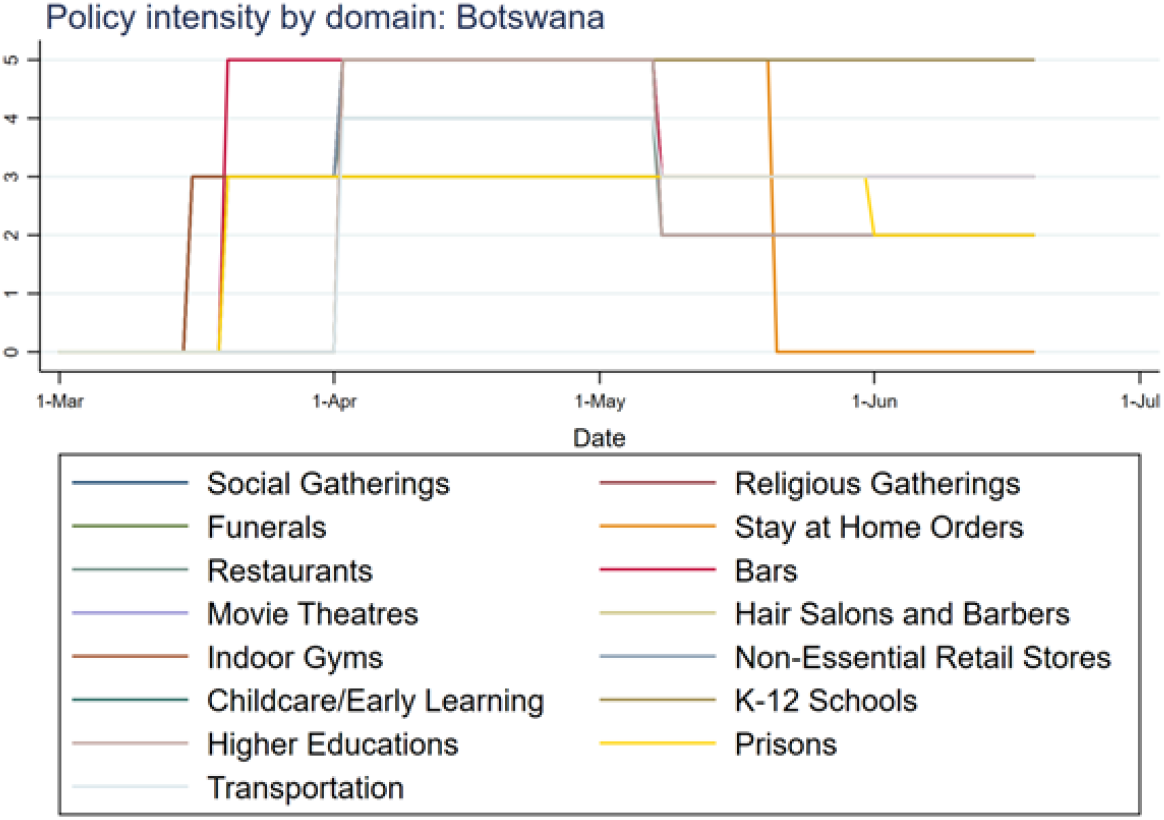
Policy domains daily scores in Botswana (other countries can be found in S1)

Figure 3 depicts the average daily physical distancing intensity across domains in the six sampled LMICs and the daily confirmed COVID-19 case data for the same period. Countries in this sample adopted mass physical distancing policies in the very early stages of the pandemic. Two countries (Botswana and Mozambique) implemented a physical distancing policy before any cases were formally documented in the country. The first case in Botswana was identified 14 days after the first policy mandate, and in Mozambique 7 days after the first policy mandate. India’s first policy mandate was implemented upon testing and confirming 119 cases, Jamaica implemented at 26 cases, Namibia at 8 cases, and the Ukraine at 1 case. Amongst the four countries that implemented policy mandates after a case was detected, the average number of days between case detection and a physical distancing mandate ranged from 10 days (Ukraine) to 47 days (India).

**Figure 3.**
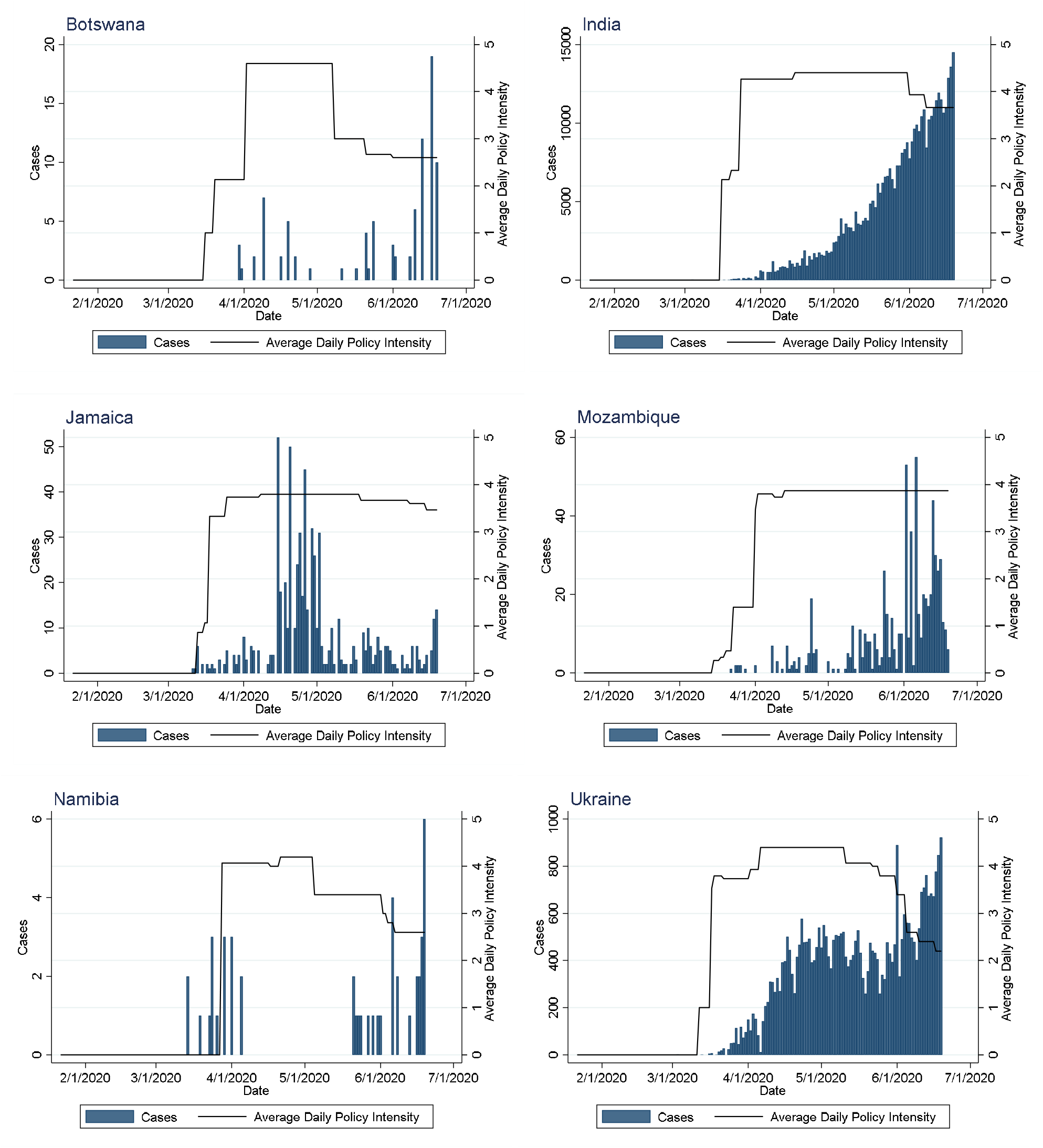
Daily policy intensity and COVID-19 cases, by country (15 domains)

### Comparing policy responses between LMICs and the U.S

Comparing the results from this analysis with the physical distancing intensity in U.S. states reveals some noteworthy differences. The results of comparing physical distancing intensity in this six LMIC sample with the U.S. (using the 14 domains common across all sample countries) varied substantially depending on which U.S. state was used for comparison. For example, the state of California had a peak intensity of 4.29, which would place California below the peak intensity for Botswana, India, and Ukraine but above Mozambique, Namibia and Jamaica. In contrast, the state of Georgia in the United States had a peak intensity of 3.07, which would place it lower than all of the LMICs in this sample. The peak intensity for the U.S. 12-state average was 3.84, which would place it lower than every LMIC in this sample except Jamaica. Figure 4 compares the average daily intensity across all seven sample countries, including the U.S. states of California, Georgia and the U.S. 12-state average.

**Figure 4.**
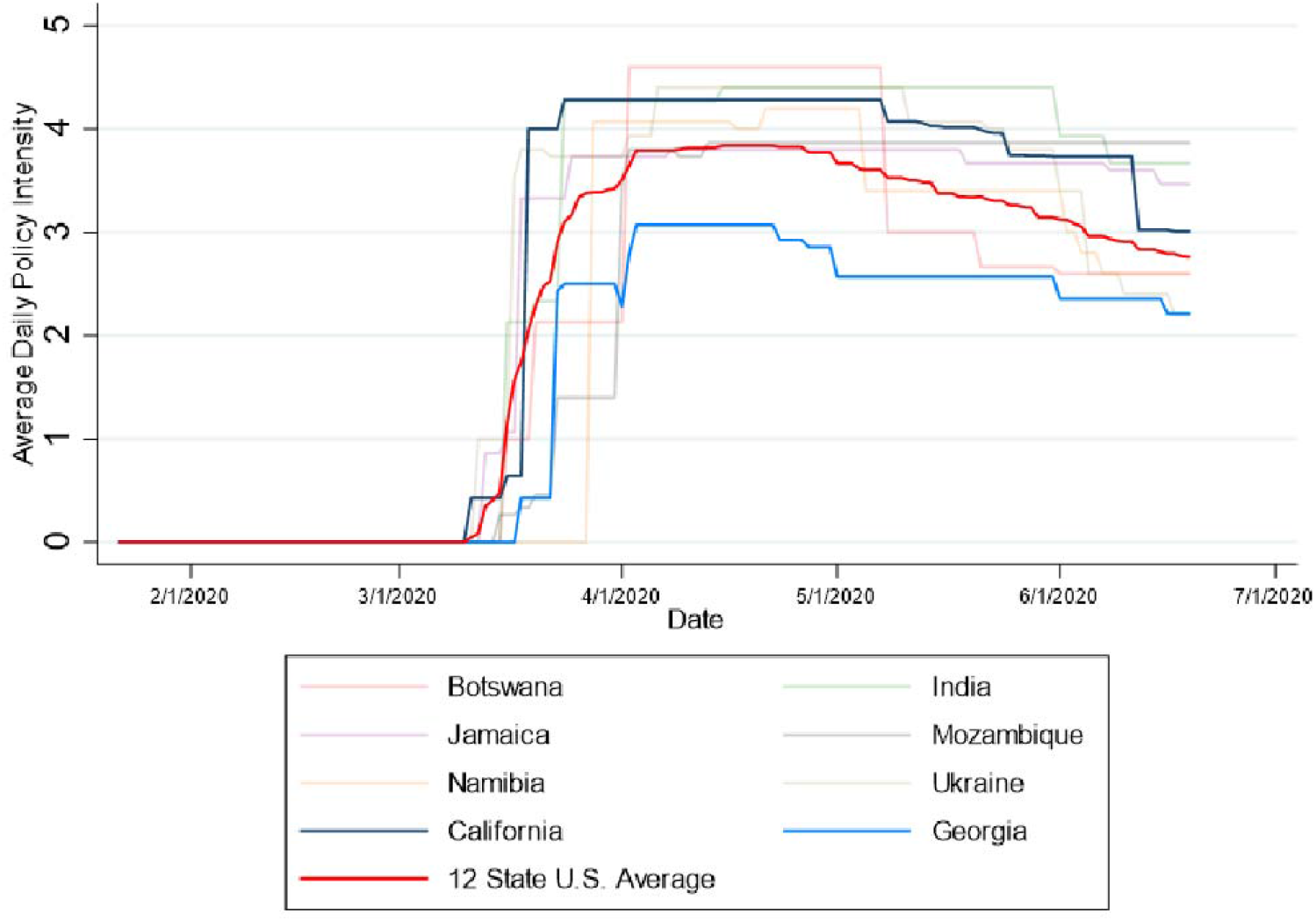
Comparison of physical distancing responses in samples LMICs and the U.S.

### Public Communication

We noted a wide variety of methods for communicating with the public regarding the degree of physical distancing recommended or required. A number of countries adopted physical distancing through Emergency Orders or other official legal instruments. These formal legal decrees require close examination to identify the nuances of what restrictions apply to which domains and in what parts of the country. This becomes increasingly difficult as decrees are amended over time. A number of countries adopted communication materials, such as infographics, targeted at the general population to communicate policies. For example, India adopted an infographic describing the key aspects of physical distancing in effect, and a red, yellow, green color-coding scheme to designate the level of physical distancing in each sub-region. Some governments used a variety of approaches to communicate with the public, including daily press briefings. For example, the governments of Botswana and Namibia provided daily TV updates, which were also streamed live online. Botswana published a regular COVID-19 task force bulletin.(16) Simplified and easy to understand communication strategies will be increasingly important if countries continue to adjust physical distancing approaches in response to changing COVID-19 epidemiological circumstances.

## Discussion

In the first 100 days of the COVID-19 pandemic, physical distancing policies adopted in the LMICs in our sample varied by timing, intensity, and scope. These findings shed light on how countries have responded to COVID-19 to date and how policies might continue to evolve over the course of the pandemic. Our primary findings were that while each LMIC country adopted physical distancing mandates across at least 14 out of 15 domains, there was substantial variability in peak physical distancing at the country and domain level. The countries with the highest peak intensity were Botswana, India and Ukraine with Jamaica having the lowest intensity peak. All six of the countries in our sample imposed physical distancing mandates early in the COVID-19 outbreak in that country. Five out of the six LMICs adopted physical distancing policies that had a peak more intense than the average peak intensity reached by a sample of 12 U.S. states. Apart from the few small modifications mentioned above in the methods section (removing nursing homes and voting, and adding public transport), we found that the Physical Distancing Intensity Framework performed well for the LMIC sample countries and was sufficiently sensitive to capture longitudinal change in the physical distancing policy responses the LMIC sample countries.

### Timing of physical distancing policies

The six LMIC countries in this sample adopted mass physical distancing policies in the very early stages of the pandemic, with two countries implementing policies before any cases were confirmed in country. While this observation may be due to limited testing, and thus invalid estimates of true disease prevalence in each setting, the detected prevalence may indicate perceived risk and policy urgency. Without the presence of reliable in-country testing, countries may have looked to other evidence to inform policy decisions. Media reports from some countries reveal that national governments were closely monitoring confirmed case counts and governmental responses in neighboring countries. For example, the growth of the COVID-19 outbreak in South Africa and the government of South Africa’s response may have played a key role in influencing the timing and approach of the aggressive policy responses observed in Namibia, Botswana, and Mozambique (all three of which share a land border with South Africa).

The public statements of the WHO may also have played a role in the early implementation of physical distancing in the LMIC sample. The *WHO’s 2019 Novel Coronavirus* (*2019*-*nCov*): *Strategic Preparedness and Response Plan* dated 14 April 2020 included the following global strategic objective: “Suppress community transmission through context appropriate infection prevention and control measures, population level physical distancing measures, and appropriate and proportionate restrictions on non-essential domestic and international travel.”(17) Notably, an earlier draft of the response plan published on 3 February 2020 did not reference mass social or physical distancing, illustrating the exponential growth of COVID-19 in February and March of 2020 and the associated realization that a more aggressive global response was required.(18) By the end of March, every country in our sample had adopted its first mandatory policy imposing some level of physical distancing to mitigate COVID-19 spread.

Months later in the pandemic, reductions in policy intensity did not align with consistent reductions in disease incidence. It is likely that social and economic costs and inequitable impact on certain groups influenced changes in policy based on community responses and behaviors.(19) For example, as a result of protests and other forms of political pressure on governmental policymakers.

### Governance & policy decision-making

We observed important differences in the governance approaches among LMICs in this sample. This was most notable in differing approaches to national versus subnational authority to mandate levels of physical distancing.(20) For example, in the U.S., which is a federation of states, the national government did not adopt any mandatory policies imposing social distancing. Rather, during the period of this review (March 1-June 19) the U.S. federal government explicitly deferred to individual state governments to decide what level of physical distancing should be imposed. In contrast, in India, which also has a governance structure that grants a great deal of authority to individual states (sometimes called a semi-federal or quasi-federal state), the central government initially only published guidelines for physical distancing. For example, the guidance issued by the central government of India regarding physical distancing was initially characterized as “guidelines” and indicated that states had the authority to adapt them; however, subsequent guidance describing the approach stated: “States/UTs shall not dilute the guidelines issued under the Disaster Management Act, 2005, in any manner, and shall strictly enforce the same.”(21) In Ukraine, the country shifted to what it referred to as a modified quarantine approach under which the central government granted individual subregional governments (called Oblast governments) the authority to impose stricter physical distancing measures than required nationally. A number of Oblast governments exercised that power to impose more intense physical distancing (e.g., Kiev Oblast).

Our review also pointed to the important role of governance structures at the national level and how the interplay between those structures can influence physical distancing approaches. For example, in Mozambique, the national legislature (Assembleia da Republica) must approve an emergency decree before it goes into effect. The President of Mozambique proposed an emergency decree imposing physical distancing measures, but the legislature altered the decree proposed by the President prior to adopting it. The emergency decree became effective later that night and reports indicate that modification led to confusion regarding the scope of decree, and in particular, whether retail stores could continue to operate.(22)

### Effects of mass physical distancing on health systems

All stay at home policies in our sample allowed leaving home to receive or provide essential health care, nevertheless, major disruptions of the health services have been reported in countries in our sample. For example, while India and Namibia have shifted to dispensing more anti-HIV medications at a time, known as multi-month dispensing (e.g., shifting from 3-months of medicine to 6-months), other countries, such as Botswana, have had to reduce the number of anti-HIV antiretroviral pills per refill, because of supply chain disruptions. Blood shortages resulting from a reduction in blood donations have also been reported in multiple countries, including India and Namibia.(23, 24) Jamaica introduced home delivery of different types of medications, including anti-HIV medications.(25) Health care and public health workers have faced stigma and discrimination due to their work on the frontline fighting COVID-19. The Government of India released guidelines on Addressing Social Stigma Associated with COVID-19 to help counteract growing stigma in India.(26) A recent modeling study estimated that deaths due to HIV, tuberculosis and malaria may increase by up to 10%, 20% and 36% respectively, over the next 5 years in LMICs due to disruptions to diagnostic and treatment services (including antiretroviral therapy) and the interruption of preventative campaigns, including routine bednet distribution.(27) Physical distancing policies have also led to a dramatic shift toward telemedicine in many countries, including India and Jamaica.(28) Adopting telemedicine approaches may mitigate some disruption to health services, but not every country or region will have the information technology infrastructure required to quickly shift to telemedicine.

### Disproportionate impact on certain populations

All of the countries in our sample adopted policies restricting commercial activity through a variety of domains, such as non-essential retail or stay at home orders. Reports from the countries in this sample indicate that these restrictions are having a disproportionate impact on certain populations. Food insecurity has been reported in many of the countries included in this analysis, including Botswana.(16) Restrictions on inter-regional and cross-border travel and closure of businesses have left large numbers of people, especially migrant workers, stranded with limited means for returning to their homes.(29) For example, regional travel restrictions in India led to mass protests in some cities from stranded migrant workers.(30) The closure of the land border between Mozambique and South Africa has led to major economic disruption to people residing on the Mozambique side of the border who rely on cross-border commerce. A number of countries, including India(31) and Botswana, have reported increases in gender-based violence during physical distancing. The Botswana Gender-Based Violence Prevention and Support Centre reported that cases of gender-based violence in the city of Gaborone had spiked in April and reflected a “sharp increase compared to months before the lockdown.” (32) In contrast, the Namibian Police recorded a decrease in reported gender-based violence in Windhoek during the period of emergency and cited the total cessation of liquor sales in Namibia during the lockdown as a possible contributing factor.(33) Physical distancing may also be having a proportionate impact on lower-income populations working in the informal sector who have little savings and must continue to work every day to provide for their families. Some countries in this analysis established financial subsidies to encourage people to stay at home, such as Botswana, Namibia, and Jamaica.(34-36)

## Conclusion

Our analysis revealed some key differences and many similarities in the tactics for instituting mass physical distancing in countries spread across four continents with varying epidemiological profiles. Additional analysis over longer durations of time will help determine which of these approaches, and the intensity of their delivery, optimally balance reduction in COVID-19 transmission with associated economic, social and health system costs. Nevertheless, this analysis helps to highlight the differing paths taken by the countries in this sample and may provide lessons to other countries regarding options available for structuring physical distancing policies in response to COVID-19 and future outbreaks of respiratory infectious diseases.

## Supporting information

Supplemental Policy Coding Datasets

## Data Availability

The datasets generated in this analysis are included in the supporting file.

## Declarations

### Ethics approval and consent to participate

Not applicable. This analysis did not involve human subjects research.

### Availability of data and materials

The datasets generated in this analysis are included in the supporting file.

## Authors’ contributions

JPL - Conceptualization; Writing – Original Draft; AM - Conceptualization; Writing – Review & Editing; Formal Analysis; KB - Conceptualization; Writing – Review & Editing; AS – Investigation; Writing-Review & Editing; Formal Analysis; FV - Investigation; Writing-Review & Editing; Formal Analysis; CA - Investigation; Writing-Review & Editing; AC - Investigation; Writing-Review & Editing; NF - Investigation; Writing-Review & Editing; JL - Investigation; Writing-Review & Editing; GO - Writing-Review & Editing SM - Investigation; Writing-Review & Editing; AP - Investigation; Writing-Review & Editing; LP - Writing-Review & Editing; FM - Investigation; Writing-Review & Editing; Formal Analysis. All authors read and approved the final manuscript.

## References

1. World Health Organization. Timeline - COVID-19 [Internet]. 2020 Apr 27 [Available from: https://www.who.int/news-room/detail/27-04-2020-who-timeline---covid-19.

2. Yuan Z, Xiao Y, Dai Z, Huang J, Zhang Z, Chen Y. Modelling the effects of Wuhan’s lockdown during COVID-19, China. Bull World Health Organ. 2020;98(7):484–94.

3. World Health Organization. WHO Director-General’s opening remarks at the media briefing on COVID-19 - 13 March 2020 [Internet]. 2020 Mar 13 [Available from: https://www.who.int/dg/speeches/detail/who-director-general-s-opening-remarks-at-the-mission-briefing-on-covid-1913-march-2020.

4. Lane J, Garrison MM, Kelley J, Sarma P, Katz A. Strengthening Policy Coding Methodologies to Improve COVID-19 Disease Modeling and Policy Responses: A Proposed Coding Framework and Recommendations. medRxiv. 2020:2020.08.12.20173740.

5. World Health Organization. Countries Botswana [Internet]. 2020 [cited 2020 Sep 24]. Available from: https://www.who.int/countries/bwa.

6. World Health Organization. Countries India [Internet]. 2020 [cited 2020 Sep 24]. Available from: https://www.who.int/countries/ind/.

7. World Health Organization. Countries Jamaica [Internet]. 2020 [cited 2020 Sep 24]. Available from: https://www.who.int/countries/jam/.

8. World Health Organization. Countries Mozambique [Internet]. 2020 [cited 2020 Sep 24]. Available from: https://www.who.int/countries/moz/en/.

9. World Health Organization. Countries Namibia [Internet]. 2020 [cited 2020 Sep 24]. Available from: https://www.who.int/countries/nam/en/.

10. World Health Organization. Countries Ukraine [Internet]. 2020 [cited 2020 Sep 24]. Available from: https://www.who.int/countries/ukr/en/.

11. World Health Organization. Countries - United States of America [Internet]. 2020 [cited 2020 Sep 24]. Available from: https://www.who.int/countries/usa.

12. Lane J, Garrison MM, Kelley J, Sarma P, Katz A. Social Distancing Policy Intensity in U.S. States [distributor]. Ann Arbor, MI: Inter-university Consortium for Political and Social Research 2020 Aug 12 [Available from: https://www.openicpsr.org/openicpsr/project/120598/version/V1/view.

13. Dong E, Du H, Gardner L. An interactive web-based dashboard to track COVID-19 in real time. Lancet Infect Dis. 2020;20(5):533–4.

14. Frey A. Mozambique: Sofala releases a token 57 of 437 prisoners covered by amnesty [Internet]. Club of Mozambique; 2020 Apr 16 [cited 2020 Sep 25]. Available from: https://clubofmozambique.com/news/mozambique-sofala-releases-a-token-57-of-437-prisoners-covered-by-amnesty-watch-157861/.

15. Frey A. Mozambique: Masks to be mandatory in transport, crowded places; taxi rules eased [Internet]. Club of Mozambique; 2020 Apr 09 [Available from: https://clubofmozambique.com/news/mozambique-masks-to-be-mandatory-in-transport-crowded-places-taxi-rules-eased-157385/.

16. Presidential (COVID-19) Task Force Bulletin. Republic of Botswana; 2020 Apr 28.

17. World Health Organization. COVID-19 Strategy Update. Geneva, Switzerland; 2020 Apr 14.

18. World Health Organization. 2019 Novel Coronavirus (2019-nCoV): Strategic Preparedness and Response Plan. Geneva, Switzerland; Draft as of 3 Feb 2020.

19. Islam N, Sharp SJ, Chowell G, Shabnam S, Kawachi I, Lacey B, et al. Physical distancing interventions and incidence of coronavirus disease 2019: natural experiment in 149 countries. Bmj. 2020;370:m2743.

20. Yan B, Zhang X, Wu L, Zhu H, Chen B. Why do countries respond differently to COVID-19? A comparative study of Sweden, China, France, and Japan. American Review of Public Administration. 2020;50(6-7):762–69.

21. Government of India, Ministry of Home Affairs, Order No 40-3/2020-DM-I(A), (2020 Apr 15).

22. Hanlon J. Mozambique: Parliament Blocks Most Sensible Action as Confused Emergency Starts [Internet]. allAfrica; 2020 Apr 5 [cited 2020 Sep 25]. Available from: https://allafrica.com/stories/202004050193.html.

23. NamBTS in desperate need of blood Calling all blood donors [Internet]. Erongo.com; 2020 Apr 22 [cited 2020 Sep 25]. Available from: https://www.erongo.com.na/news/nambts-in-desperate-need-of-blood2020-04-22/.

24. Jaiswal PB. Maharashtra faces shortage of blood supply due to COVID-19 scare [Internet]. TheWeek; 2020 Mar 28 [cited 2020 Sep 25]. Available from: https://www.theweek.in/news/india/2020/03/28/maharashtra-faces-shortage-of-blood-supply-due-to-covid-19-scare.html.

25. NHF Commences Delivery of Mediation for Drug Serv Patients 65 and Older [Internet]. National Health Fund; 2020 Apr 5 [cited 2020 Sep 25]. Available from: https://www.nhf.org.jm/news/item/nhf-commences-delivery-of-medication-for-drug-serv-patients-65-and-older.

26. Government of India, Ministry of Health and Family Welfare, Addressing Social Stigma Associated with COVID-19. 2020.

27. Hogan AB, Jewell BL, Sherrard-Smith E, Vesga JF, Watson OJ, Whittaker C, et al. Potential impact of the COVID-19 pandemic on HIV, tuberculosis, and malaria in low-income and middle-income countries: a modelling study. Lancet Glob Health. 2020;8(9):e1132–e41.

28. Murphy J. COVID Drives Patients to Log On To Online Doctors [Internet]. The Gleaner; 2020 Mar 17 [cited 2020 Sep 25]. Available from: http://jamaica-gleaner.com/article/lead-stories/20200317/covid-drives-patients-log-online-doctors.

29. Rao M. Over 10 agonizing days, this migrant worker walked and hitchhiked 1,250 miles home. India’s lockdown left him no choice [Internet]. CNN; 2020 May 31 [cited 2020 25 Sep]. Available from: https://www.cnn.com/2020/05/30/asia/india-migrant-journey-intl-hnk/index.html.

30. Shantha S. ‘Let us Go Home’: No Sign of Relief in PM’s Speech, Migrant Workers Take to Mumbai Streets [Internet]. The Wire; 2020 Apr 14 [cited 2020 Sep 25]. Available from: https://thewire.in/labour/mumbai-bandra-migrant-covid-19.

31. Ratnam D. Domestic violence during COVID-19 lockdown emerges as serious concern [Internet]. Hindustan Times; 2020 Apr 26 [cited 2020 Sep 25]. Available from: https://www.hindustantimes.com/india-news/domestic-violence-during-covid-19-lockdown-emerges-as-serious-concern/story-mMRq3NnnFvOehgLOOPpe8J.html.

32. Gaborone Records 114 Gender Based Violence Cases During Lockdown [Internet]. The Botswana Gazzette; 2020 Apr 27 [cited 2020 Sep 25]. Available from: https://www.thegazette.news/news/gaborone-records-114-gender-based-violence-cases-during-lockdown/31125/#.X242j2hTmUk.

33. Wolf V. Fewer GBV cases reported during lockdown [Internet]. The Namibian; 2020 Jul 09 [cited 2020 Sep 25]. Available from: https://www.namibian.com.na/202471/archive-read/Fewer-GBV-cases-reported-during-lockdown.

34. Republic of Botswana, Presidential (COVID-19) Task Force Bulletin. 2020 Apr 14.

35. Ngatjiheue C. Govt rolls out N$8,1b COVID-19 stimulus [Internet]. The Namibian; 2020 Apr 02 [cited 2020 Sep 25]. Available from: https://www.namibian.com.na/199723/archive-read/Govt-rolls-out-N$81b-Covid-19-stimulus.

36. Jamaica Ramps Up Social and Economic Support in COVID-19 Response [Internet]. International Monetary Fund; 2020 May 28 [cited 2020 Sep 25]. Available from: https://www.imf.org/en/News/Articles/2020/05/27/na052720-jamaica-ramps-up-social-and-economic-support-in-covid-19-response.

